# Impact of Transport Modality on Time to Endovascular Thrombectomy

**DOI:** 10.1101/2025.11.19.25340623

**Authors:** Knut Taxbro, Avan Sabir Rashid, Gabriel Skallsjö, Per Arnell, Karl Chevalley, Alexandros Rentzos, Rianne Goselink

## Abstract

**Background and Purpose:** Endovascular thrombectomy (EVT) is a time-critical treatment for acute ischemic stroke; however, timely access to comprehensive stroke centers (CSCs) is often limited by distance. Regional variations in prehospital transport strategies, particularly Helicopter Emergency Medical Services (HEMS) use, exist in Sweden, which may contribute to inequities in care. This study analyzed how transport modality affected the time to EVT for rural patients across two large Swedish regions with differing transport strategies.

**Methods:** A retrospective registry-based study was performed, using comprehensive stroke registries and ambulance records from 2018 to 2022. All patients who underwent EVT in two healthcare regions (Västra Götaland and Southeastern healthcare regions) were included. The primary analysis focused on those located ≥50 km from the CSC. The main outcome was the time from emergency medical service (EMS) dispatching to EVT start. Logistic regression was used to assess the odds of receiving EVT within 180 minutes.

**Results:** Among the 1,222 patients, 623 (51%) were ≥50 km from a CSC. Direct CSC transfer and HEMS use were more frequent in Västra Götaland compared to the Southeastern region (52.0% vs. 28.4% and 8.9% vs. 0.9%, respectively; P<0.001 and P=0.003, respectively). For patients ≥50 km away, HEMS transport yielded a shorter median dispatch-to-EVT time than ground transport (224.2 vs 287.5 min; P<0.001). After adjusting for distance, HEMS was associated with a 3.6-fold higher likelihood of EVT within 180 min (OR 3.6 [95% CI 1.6-7.8], P=0.001).

**Conclusions:** Geographical distance significantly delays time to EVT. Use of HEMS markedly shortens transport time leading to timelier EVT for patients with long distances to CSC and has the potential to mitigate regional disparities. Integrating HEMS into stroke transport protocols is essential to ensure timely and equitable EVT access for rural patients.

## Introduction

Acute ischemic stroke (AIS) represents a significant global health burden and is a leading cause of death and the primary cause of long-term disability. (1) Among the various stroke types, large vessel occlusion (LVO) strokes are particularly severe, accounting for a substantial proportion of disabling strokes and often leading to profound neurological deficits. In Sweden, with a population of 10.7 million, approximately 25,500 individuals suffer a stroke each year (238 per 100.000 inhabitants), with a point prevalence of 120,000 people living with the long-term consequences of stroke. (2) Interventions during the acute phase aim to minimize the time from symptom onset to diagnosis and treatment. The acute care pathway involves healthcare resources both outside and within hospitals. The out-of-hospital chain of care for stroke patients comprises the allocation and execution of pre-hospital transport for definite care.

Several multicenter, open-label randomized trials have demonstrated that early Endovascular Thrombectomy (EVT) is safe and effective in reducing the disease burden following ischemic stroke. (3–7) EVT in combination with intravenous thrombolysis (IVT) is superior to IVT alone in patients with confirmed anterior cerebral circulation occlusion. (8) The number needed to treat (NNT) ranges from 3 to 7.5 for one additional patient to achieve functional independence. (3–7, 9) Not only does the effectiveness of treatment decline rapidly with time after stroke onset, but there are also upper time limits beyond which treatment is no longer effective. Therefore, reducing the time from symptom onset to diagnosis and treatment in stroke care is a critical task that requires well-structured care processes and efficient transport systems to achieve optimal patient outcomes.

Sweden has an area of approximately 450-thousand square kilometers, and EVT is available at seven Comprehensive Stroke Centers (CSC), creating logistical challenges towards bringing potential EVT candidates to the appropriate level of care within the given time limits. To mitigate the effect of long transfer times, some Swedish regions have prioritized the utilization of helicopter emergency medical services (HEMS) for patients with stroke, whereas others have not. In addition, a report put forward by the National Quality Registry points out that access to specialized stroke care, particularly advanced treatments such as thrombectomy, is not equally distributed geographically. (10) Internationally, evidence remains inconclusive as to whether direct transport to CSC and HEMS is associated with improved health outcomes.

In this study, we will include patients from two regions (Västra Götaland Region [VGR] and the South-Eastern Healthcare Region [SÖSR]) including nearly 1/3 of the population of Sweden. Although direct transport to the CSC by ground emergency medical services (GEMS) or helicopter emergency medical services (HEMS) is a possible approach for patients who meet the criteria for potential mechanical thrombectomy, SÖSR does not have the potential for direct triage to thrombectomy. In SÖSR, all patients with stroke are initially transported to the nearest primary stroke center (PSC) for computed tomography (CT) and intravenous thrombolysis, after which secondary ground EMS transport from the PSC to the CSC is arranged if indicated. Both regions have a substantial proportion of inhabitants in rural areas, as far as 350 km from the CSC.

This study aimed to analyze transport logistics among stroke patients who underwent EVT across two catchment areas. Particular focus will be placed on patients in rural areas who suffer a stroke of 50 km or more from a CSC, and on the effect of transport to CSC by either ground or helicopter transport.

## Methods

A retrospective, registry-based observational study was performed, including patients with stroke prompting EVT between 1^st^ of January 2018 and the 31^st^ of December 2022. The data were sourced from the National Quality Registry (RiksStroke) and ambulance records. This study aimed to examine the time to EVT from the initial medical emergency service dispatch among patients ≥ 50 km from CSC.

### Data availability statement

Data collected for the study, including individual participant data and a data dictionary defining each field in the set, were registry-based and derived from multiple cross-matched registry sources; therefore, we were unable to forward this to others due to registry regulations.

### Ambulance Resources and Triage Routines

In the VGR, patients within a 45-minute transport radius of Sahlgrenska University Hospital (SUH) could be transported directly if the modified National Institutes of Health Stroke Scale (mNIHSS) score was ≥ 6, which can be at the CSC within 6 h from the last-seen-well time (for the detailed triage decision matrix, see Supplement 1). All others were first taken to the local nearest PSC for radiological imaging and, if indicated, treatment with IVT. In the SÖSR, there was no direct-to-CSC triage routine, given certain time limits during the study period.

In VGR, patients have access to helicopter transfer, either directly to the CSC or PSC from the point-of-pickup (PUP), or as an interfacility transfer (IFT) between the PSC and CSC. The helicopter is a twin engine AW169 with a cruise speed of 140 knots/260 km per hour. The crew consists of four (nurse, physician, and two pilots) and is operational 24 hours a day. When HEMS is tasked, hospital stroke physicians, ground ambulance resources and helicopter emergency medical services frequently coordinate and collaborate to facilitate rapid transport to definitive care. This often involves rendezvous operations in which the helicopter meets the ground ambulance along the transport route for patient transfer. Structured handovers are conducted *en route* to minimize delays and optimize the treatment timelines. All HEMS tasks must be approved by the on-duty HEMS physician and have a perceived time benefit over the ground transport. Prehospital transport logistics are likely to be more complex if the patient is far from definitive care; therefore, the study placed particular emphasis on patients who experienced stroke symptoms 50 km or more from a CSC and were treated with EVT within 10 hours from the initial EMS dispatch. Distance is preferred over time as a metric because it can be more precisely defined using the geolocated point where ambulance services attend to the patient.

### Geographical and Demographic Characteristics

VGR has a population of 1,752,072, an area of 23,800 km², and a population density of 73.6 inhabitants per km². The longest road distance to CSC was 212 km (Gullspång to Gothenburg). SÖSR has a population of 1,086,183, an area of 32,161 km², and a population density of 33.8 inhabitants per km², with the longest road distance to the CSC being 321 km (Byxelkrok to Linköping). (11)

### Definitions

PSC refers to a hospital that receives patients with stroke for computer-assisted tomography (CT) and IVT. A comprehensive stroke center (CSC) is a hospital with EVT capabilities. PUP refers to the geolocated point where an ambulance first loads a patient who later undergoes EVT. The time measured from the initial EMS dispatch to the start of the EVT procedure (known as groin time) defined the time to EVT.

### Inclusion and Exclusion Criteria

Patients eligible for inclusion were aged > 18 years with a diagnosis of stroke (International Classification of Disease 10 code I63) and who had undergone EVT between 2018 and 2022. Exclusion criteria included cases in which time and geolocation data required for the primary endpoint were missing, and patients not managed by ambulance services. To mirror the most time-critical cases, EVT performed within 10 h (600 min) of the initial ambulance dispatch was included in the final analysis.

### Outcome Measures

The primary outcome was the time from emergency dispatch to EVT for patients located ≥ 50 km from the CSC. Secondary outcomes included time from emergency dispatch to arrival at CSC, the proportion of patients directly triaged to CSC, mode of transport (GEMS or HEMS), and predictors (distance from PUP to CSC and mode of transport) likely to influence time to EVT using a logistic regression model. In these models, the dependent variable was EVT within 3 h and 6 h from the initial EMS dispatch and was analyzed separately.

### Data Management

Primary data were obtained from the Swedish Stroke Registry. Geolocations and dispatch times were retrieved from ambulance records (Ambulink and Paratus), which required data linkage between ambulance records and Riksstroke using unique Swedish personal identification numbers. Data linkage between ambulance records and primary data was performed by a Riksstroke statistician. Geodesic distances in kilometers were calculated in Microsoft Excel® using the following function: =ACOS(COS(RADIANS(90-LAT1))*COS(RADIANS(90-LAT2))+SIN(RADIANS(90-LAT1))*SIN(RADIANS(90-LAT2))*COS(RADIANS(LONG1-LONG2)))*6371. Driving distances were retrieved using the Google Maps function in Microsoft Excel. All geolocation coordinates were in World Geodetic System 1984 (WGS 84) format. Data were compiled in a digital database (Microsoft Excel®) and subsequently exported in coded form to a statistical software for analysis (SPSS v. 29 IBM Armonk, NY, USA). The database will be stored as a medical record for 10 years following the data collection. Endnote PaperPal® was used for proofreading and linguistic improvements.

### Ethics statement

The study protocol was approved by the Swedish Ethical Review Authority (Dnr 2023-00856-01) and was conducted in accordance with the Declaration of Helsinki. Owing to the retrospective observational design of the study, informed consent was not required.

### Statistical Analysis

Continuous variables were reported as median with 25th-75th percentiles expressed as interquartile ranges (IQR), while categorical variables were summarized as counts and percentages. Comparisons between groups were performed using Chi2-, Fisher, or Mann-Whitney tests, as appropriate. All p-values were two-tailed, and statistical significance was set at P<0.05. Results from the regression analyses are presented as odds ratios (OR) with 95% confidence intervals (CI) and p values. Statistical analyses were performed using SPSS version 29 (IBM, Armonk, NY, USA).

## Results

A total of 1335 unique EVTs were performed between 2018 and 2022 across the catchment area of which complete data was available in 1222 EVT procedures. Of these, 623 (51%) were 50 km or more from CSC (Figure 1).

**Figure 1.**
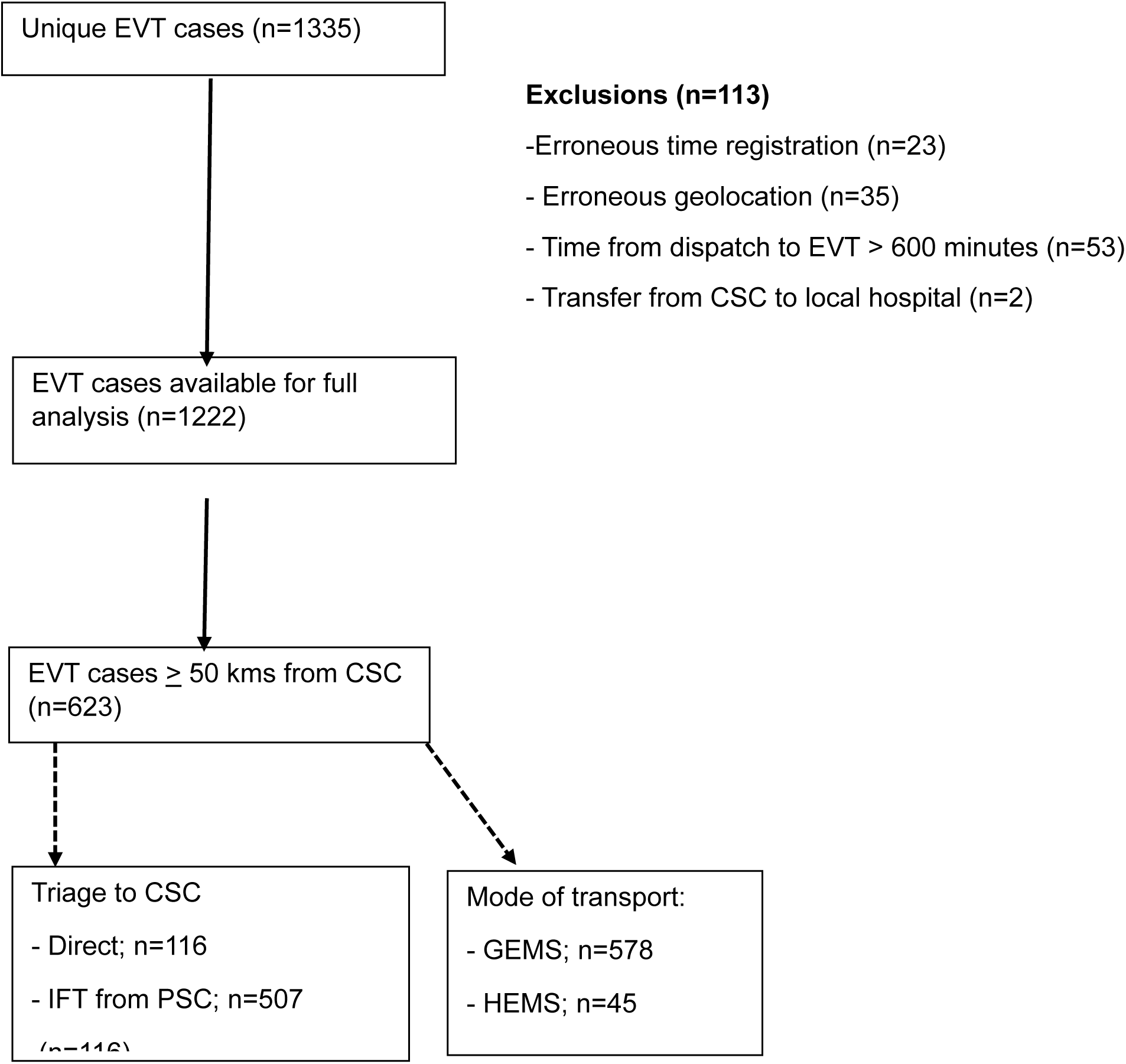
Study outline. EVT;endovascular thrombectomy, CSC;comprehensive stroke centre, GEMS;ground emergency medical service, HEMS;helicopter emergency medical service, IFT;interfacility transfer.

### Patient Characteristics and Geographical Distribution

Table 1 summarizes the baseline characteristics of the patients across the two catchment areas: VGR and SÖSR. The median age of the patients in both the VGR and SÖSR groups was 77 years (IQR 69-84 for VGR, 68-83 for SÖSR), with no significant difference observed (P=0.620). Sex distribution was also similar between regions, with 49% females in the VGR and 48% in the SÖSR (P=0.869). The level of consciousness at hospital arrival showed a trend towards more obtunded or unconscious patients in the SÖSR group, although the difference was not statistically significant (P=0.058). Other characteristics, such as previous stroke, atrial fibrillation, active smoking, hypertension, and diabetes, were comparable between regions.

**Table 1.**
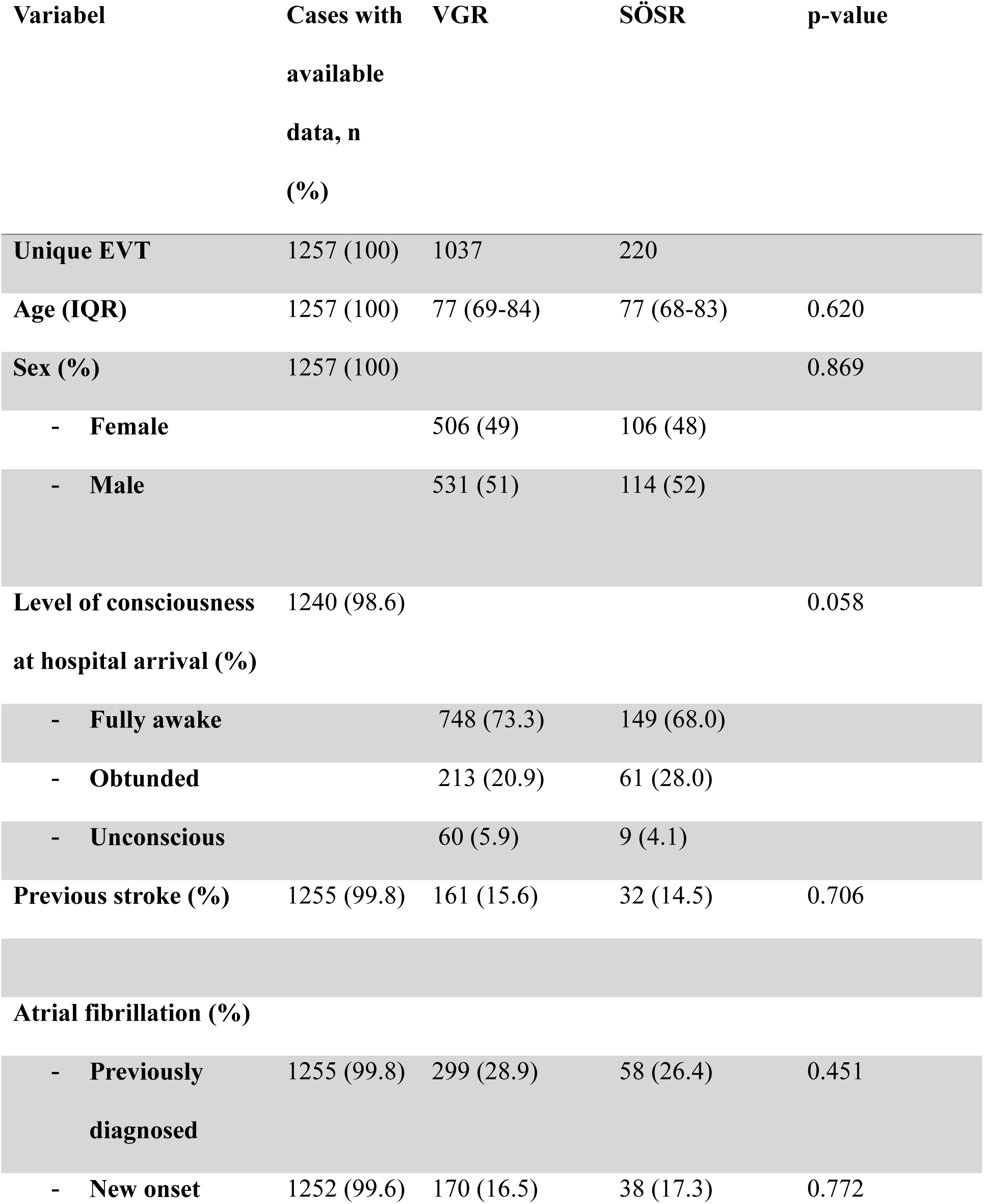
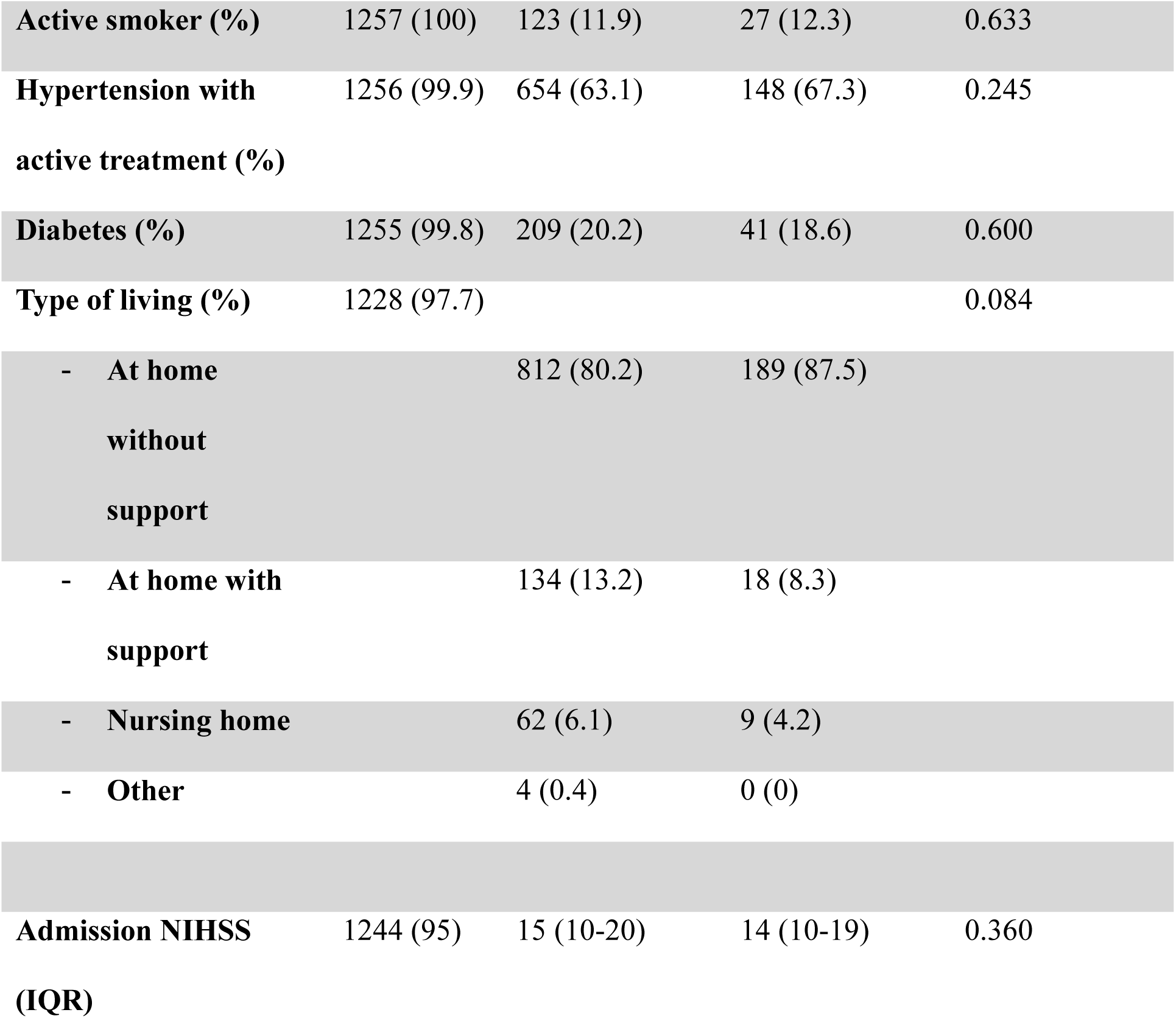
Patient characteristics at baseline across the two catchment areas. Mechanical thrombectomy; EVT. NIHSS; National Institute of Health Stroke Scale. IQR; interquartile range.

| **Variabel** | **Cases with**<br>**available**<br>**data, n**<br>**(%)** | **VGR** | **SÖSR** | **p-value** |
| --- | --- | --- | --- | --- |
| **Unique EVT** | 1257 (100) | 1037 | 220 |  |
| **Age (IQR)** | 1257 (100) | 77 (69-84) | 77 (68-83) | 0.620 |
| **Sex (%)** | 1257 (100) |  |  | 0.869 |
| - Female |  | 506 (49) | 106 (48) |  |
| - Male |  | 531 (51) | 114 (52) |  |
| **Level of consciousness**<br>**at hospital arrival (%)** | 1240 (98.6) |  |  | 0.058 |
| - Fully awake |  | 748 (73.3) | 149 (68.0) |  |
| - Obtunded |  | 213 (20.9) | 61 (28.0) |  |
| - Unconscious |  | 60 (5.9) | 9 (4.1) |  |
| **Previous stroke (%)** | 1255 (99.8) | 161 (15.6) | 32 (14.5) | 0.706 |
| **Atrial fibrillation (%)** |  |  |  |  |
| - Previously<br>diagnosed | 1255 (99.8) | 299 (28.9) | 58 (26.4) | 0.451 |
| - New onset | 1252 (99.6) | 170 (16.5) | 38 (17.3) | 0.772 |
| **Active smoker (%)** | 1257 (100) | 123 (11.9) | 27 (12.3) | 0.633 |
| **Hypertension with active treatment (%)** | 1256 (99.9) | 654 (63.1) | 148 (67.3) | 0.245 |
| **Diabetes (%)** | 1255 (99.8) | 209 (20.2) | 41 (18.6) | 0.600 |
| **Type of living (%)** | 1228 (97.7) |  |  | 0.084 |
| - At home without support |  | 812 (80.2) | 189 (87.5) |  |
| - At home with support |  | 134 (13.2) | 18 (8.3) |  |
| - Nursing home |  | 62 (6.1) | 9 (4.2) |  |
| - Other |  | 4 (0.4) | 0 (0) |  |
| **Admission NIHSS (IQR)** | 1244 (95) | 15 (10-20) | 14 (10-19) | 0.360 |

Regarding the geographical distribution, Table 2 illustrates the geodesic and driving distances from PUP to CSC. There was a significant difference in the distribution of both geodesic and driving distances between VGR and SÖSR (P<0.001 for both). A higher proportion of patients in the SÖSR group were located further away from definitive care (CSC) than those in the VGR group. For instance, 13.4% of SÖSR patients had a geodesic distance of 150-199 km to the CSC compared to 5.0% in the VGR. Similarly, 10.4% of the SÖSR patients had a driving distance of 200-249 km, whereas only 1.3% of the VGR patients were in this range. Approximately half of the patients in both regions were located 50 km or more from the CSC (49.4% in the VGR and 55.0% in the SÖSR; P=0.202) (Figure 3).

**Table 2.** Distances and modes of transport across two catchment areas. EVT;endovascular thrombectomy, CSC;comprehensive stroke centre, GEMS;ground emergency medical service, HEMS;helicopter emergency medical service.

|  | Cases with<br>available<br>data, n (%) | VGR | SÖSR | p-value |
| --- | --- | --- | --- | --- |
| **Geodesic distance**<br>**between Point-of-**<br>**Pickup and CSC (%)** | 1224 (100) | 1007 (100) | 217 (100) | <0.001 |
| **0 km – 49 km** |  | 581 (57.7) | 116 (53.5) |  |
| **50 km - 99 km** |  | 276 (27.4) | 30 (13.8) |  |
| **100 km – 149 km** |  | 100 (9.9) | 33 (15.2) |  |
| **150 km – 199 km** |  | 50 (5.0) | 29 (13.4) |  |
| **200 km – 249 km** |  | 0 (0) | 9 (4.1) |  |
| **Driving distance**<br>**between Point-of-**<br>**Pickup and CSC (%)** | 1222 (100) | 1010 (100) | 212 (100) | <0.001 |
| **0 km – 49 km** |  | 504 (49.9) | 99 (44.8) |  |
| **50 km - 99 km** |  | 288 (28.5) | 32 (15.1) |  |
| **100 km – 149 km** |  | 118 (11.7) | 33 (15.6) |  |
| **150 km – 199 km** |  | 87 (8.6) | 21 (9.9) |  |
| **200 km – 249 km** |  | 13 (1.3) | 22 (10.4) |  |
| **≥250 km** |  | 0 (0) | 9 (4.2) |  |
| **Point-of-Pickup $\geq 50$ km from CSC** | 623 (51.0) | 507 (49.4) | 116 (55.0) | 0.202 |
| **Direct transfer to CSC (%)** | 1222 (100) | 526 (52.0) | 60 (28.4) | <0.001 |
| **Direct transfer to CSC when Point-of-Pickup $\geq 50$ km from CSC (%)** | 623 (51.0) | 110 (20.6) | 6 (4.8) | <0.001 |
| **Mode of transport to CSC if Point-of-Pickup $\geq 50$ km from CSC (%)** |  | 507 (100) | 116 (100) | 0.003 |
| HEMS (%) |  | 45 (8.9) | 1 (0.9) |  |
| GEMS (%) |  | 463 (91.1) | 115 (99.1) |  |

### Transport Logistics and Time to Treatment

Direct transfer to a CSC was significantly more common in the VGR group (52.0%) than in the SÖSR group (28.4%) (P<0.001). When considering patients whose PUP was > 50 km from the CSC, direct transfer to the CSC was observed in 20.6% of cases in VGR versus 4.8% in SÖSR (P<0.001). The mode of transport to CSC also differed significantly between the regions (P=0.003). HEMS transport was utilized by 8.9% of the patients in the VGR, whereas only 0.9% of the patients in the SÖSR were transported by HEMS.

Table 3 presents a comparison of the time from dispatch to CSC arrival and the time from dispatch to EVT across different modes of transport for patients situated 50 km or more from the CSC. HEMS transport was associated with significantly shorter times than GEMS transport was. The median dispatch to CSC-arrival time for HEMS was 188.1 minutes (IQR 148.5-212.7) compared to 254.3 minutes (IQR 200.1-311.2) for GEMS (P<0.001). Similarly, the median dispatch to EVT time was 224.2 minutes (IQR 183.1-238.7) for HEMS versus 287.5 minutes (IQR 236.8-338.9) for GEMS (P<0.001), as visualized in Figure 2.

**Figure 2.**
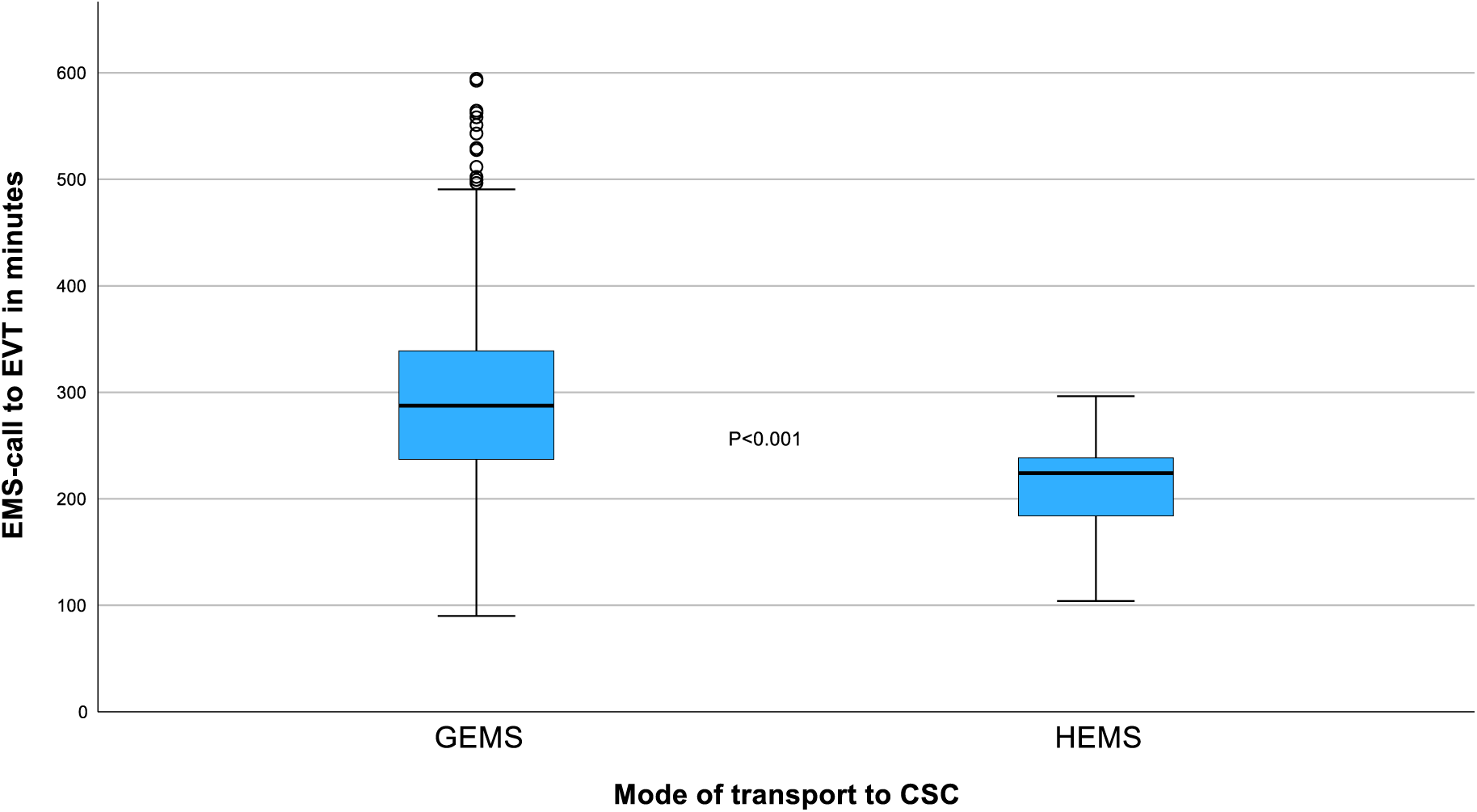
Box-plot visualizing the time in minutes between first EMS dispatch to mechanical thrombectomy in patients 50 kilometres or more from the treatment facility. EVT;endovascular thrombectomy, CSC;comprehensive stroke centre, GEMS;ground emergency medical service, HEMS;helicopter emergency medical service.

**Figure 3.**
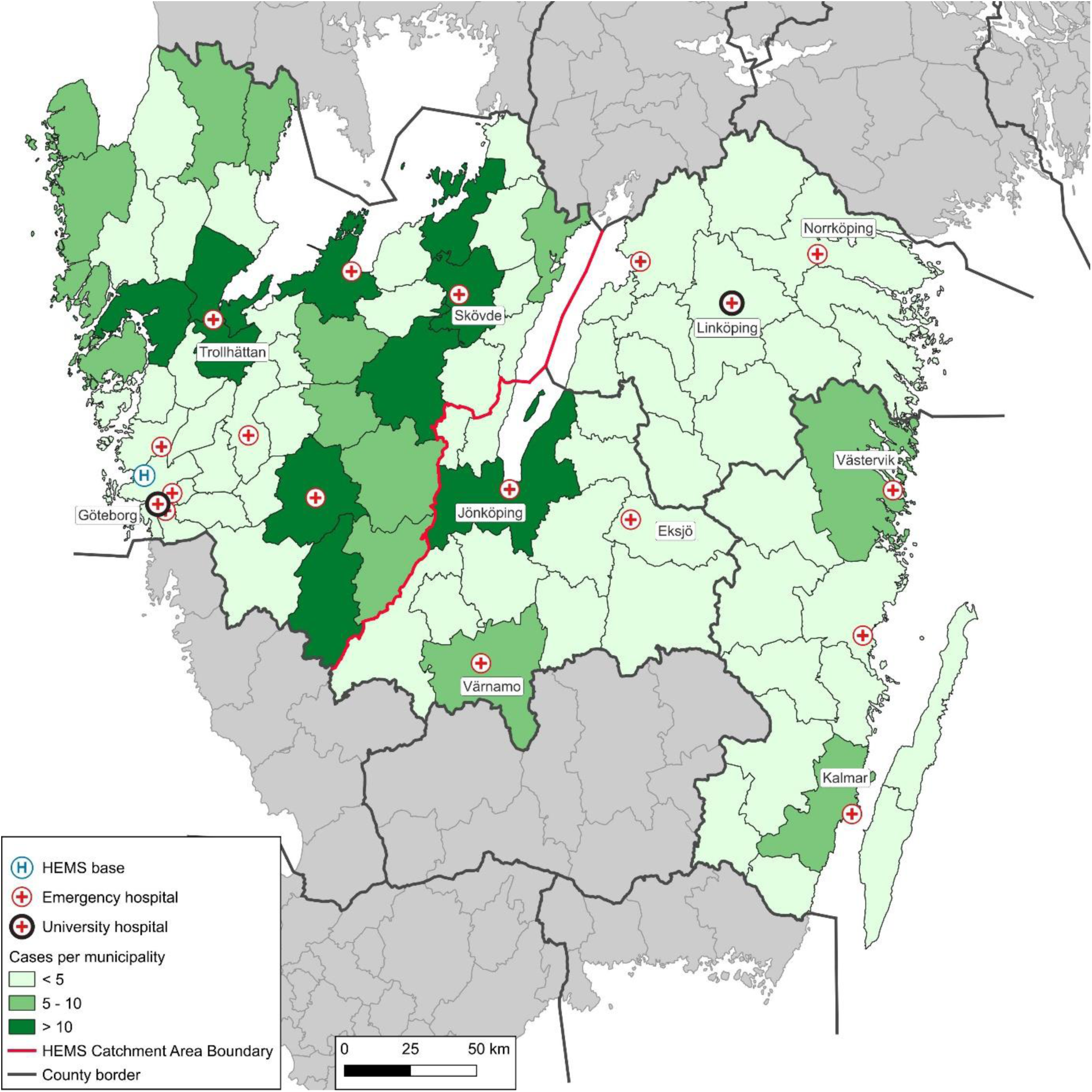
Study catchment area in color. Only cases with pick-up point > 50 km from a comprehensive stroke center are included. HEMS: Helicopter Emergency Helicopter Service. University hospitals have endovascular thrombectomy capacity.

**Table 3.** Comparison of distances between pickup point and CSC, and of time from first dispatch to CSC arrival and EVT across modes of transport in patients situated 50 kilometres or more from the CSC. 623 patients were included in the analysis. Both primary and interfacility missions are included. EVT;endovascular thrombectomy, CSC;comprehensive stroke centre, GEMS;ground emergency medical service, HEMS;helicopter emergency medical service, IFT;interfacility transfer. IQR;interquartile range.

|  | GEMS transport | HEMS transport | p-value |
| --- | --- | --- | --- |
| **Geodesic distance from initial point-of-pickup to CSC, km (IQR)** | 77.4 (59.9-116.5) | 114.8 (74.8-138.8) | 0.188 |
| **Driving distance from pick-up-point directly to CSC, km** | 97.5 (59.9-116.5) | 139.0 (76.7-138.8) | 0.051 |
| **Dispatch to CSC- arrival, minutes (IQR)** | 249.6 (196.6-308.9) | 180.0 (144.1-210.9) | <0.001 |
| **Dispatch to EVT, minutes (IQR)** | 287.5 (236.8-338.9) | 224.2 (183.1-238.7) | <0.001 |

After controlling distance, patients transported by HEMS had 3.6 times higher odds of receiving EVT within 180 minutes than those transported by GEMS (OR 3.6 [95% CI 1.6-7.8], P=0.001). Distance to EVT emerged as a strong and significant predictor of whether treatment was received within 360 minutes, and for every one-kilometer increase in distance from the CSC, the odds of receiving EVT decreased by 1% (OR 0.99 [95% CI 0.98-0.99], p<0.001. Very long distances (> 250 km) did not show a significant difference, which was likely due to the very small sample size (n=9). All patients with HEMS received treatment within 360 minutes, preventing reliable interpretation of HEMS as a predictor of treatment within 600 minutes in the logistic regression model.

## Discussion

### Main Findings

This study highlights a critical challenge in the delivery of timely EVT for acute ischemic stroke for patients in rural areas where a substantial proportion of patients experience stroke symptoms at long distances from CSCs. In this study, we showed that patients in rural locations less frequently receive timely EVT and that the use of HEMS can minimize this time delay. Patients transported by HEMS received treatment significantly faster, with an OR of 3.6 for receiving EVT within 180 minutes of the initial emergency medical services (EMS) dispatch, compared to those transported by ground.

### Results in Context

The observed regional differences in direct transfers to CSCs and the utilization of HEMS likely reflect variations in the local stroke care protocols. In regions characterized by long distances and the limited availability of HEMS, clinical strategies must prioritize rapid diagnosis and thrombolysis at the nearest PSC, followed by IFT for EVT. While this approach may be pragmatic, it raises concerns regarding health equity regardless of geographical location, as expressed in Swedish healthcare legislation. Regional healthcare authorities’ autonomy contributes to inconsistencies in prioritization and resource allocation, potentially disadvantaging patients in remote areas. Access to HEMS has emerged as a pivotal enabler of equitable access to EVT, offering geographically distant patients a chance at outcomes comparable to those of patients residing near CSCs. (12–14) Although cost is often cited as a barrier to broader HEMS implementation, recent studies, including the comprehensive work by Ennab-Vogel et al., suggest that, after considering the positive health impact of timely access to EVT following acute ischemic stroke, HEMS is a cost-effective solution for stroke transport. In this model, the most cost-effective solution would involve both shortening the geographical distance and increasing the speed of travel between patients and thrombectomy services by increasing the number of HEMS bases as well as expanding the number of EVT-capable centers in Sweden. This solution proposed an average incremental net monetary benefit of €11,050 per patient, reflecting the substantial health and economic benefits. (15) Furthermore, patients must be transported to the geographically closest appropriate stroke center regardless of administrative health region boundaries. This “closest center” principle would dismantle logistical barriers and ensure that speed of access, not regional affiliation, dictates patient care. In this context, a new government inquiry into the state control of air ambulance operations is proposed, presenting a vital opportunity to address these service gaps. For this initiative to be credible, it must lead to improved national coverage and guarantee universal access to helicopter ambulances, truly delivering equal care to everyone in the country.

The optimal prehospital transport strategy for stroke patients, either direct transport to a CSC or initial stabilization at a PSC, is still under debate. Conflicting evidence, such as from the RACECAT trial, often complicates this picture. (16) However, the applicability of such trials to Scandinavian geography with vast distances and few CSCs is questionable.

A more productive approach is to move past this general debate and to focus on clear clinical scenarios. Given the overwhelming evidence that intravenous thrombolysis should not be skipped prior to thrombectomy when a patient is eligible, the discussion should center on identifying patients who unequivocally benefit from the direct-to-CSC model. (8)Two such groups were evident. First, patients with contraindications to thrombolysis require direct access to EVT. Second, patients for whom direct transport to the CSC is demonstrably faster than the entire PCS first, followed by the CSC process, thereby minimizing time-to-treatment and avoiding secondary transfer. An exception is when the PSC lies on the direct transport route to the CSC, which makes it an efficient initial stop.

In some regions of the world, patients with stroke symptoms can be treated with dedicated mobile stroke ambulances capable of radiological imaging and thrombolytic treatment, whenever appropriate. This strategy has shown promising results and enables rapid diagnosis and treatment. (17) However, while these highly specialized ambulances originated and were initially extensively tested in urban areas, their cost-effectiveness has yet to be demonstrated in rural areas, although some data support their use for distances of up to 250 km using rendezvous approaches with regular ambulances. (17)

### Strengths and Limitations

A key strength of this study is its focus on the logistical challenges faced by stroke patients located far from CSCs, an often-underexplored aspect of stroke care. The use of contemporary data and accurate geolocation and time metrics from ambulance medical records enhances the reliability of our findings. However, this study has several limitations that must be acknowledged. This study did not report patient-centered outcomes, although the time from symptom onset to treatment was closely linked to clinical outcomes. Additionally, we were unable to differentiate between anterior and posterior circulation strokes and provide data on “false positive” (LVO-mimics)—patients transported to CSCs without an EVT indication.

## Conclusion

Timely access to EVT was significantly influenced by the distance from the CSCs. HEMS is a cost-effective option with the potential to overcome delays associated with geographic barriers and should be considered a critical component of equitable stroke care. Ensuring HEMS availability across all regions is essential to uphold the principles of health equity and optimize outcomes for all stroke patients.

## Data Availability

The data supporting the findings of this study contain confidential and proprietary institutional information and/or protected participant data; therefore, they are not publicly available and cannot be shared.

## Acknowledgments

For their invaluable help throughout the project.

**Åke Holmberg**, registered nurse/register coordinator, Emergency and Vascular Access Service (EVAS), Skånes Universitetssjukhus, Lund.

**Zlatko Mujanovic**, Department of Anesthesia and Intensive Care Medicine, Ryhov County Hospital, Jönköping, Sweden.

**Magnus Nilsson**, Development Officer, Region Västra Götaland, Gothenburg, Sweden.

## List of abbreviations

AIS: Acute Ischemic Stroke
CI: Confidence Intervals
CSC: Comprehensive Stroke Centre
CT: Computer Assisted Tomography
EMS: Emergency Medical Services
EVT: Endovascular Thrombectomy
GEMS: Ground Emergency Medical Services
HEMS: Helicopter Emergency Medical Services
IFT: Interfacility Transfer
IQR: Interquartile Range
IVT: Intravenous Thrombolysis
LVO: Large Vessel Occlusion
mNIHSS: modified National Institutes of Health Stroke Scale
NNT: Number Needed to Treat
OR: Odds Ratio
PSC: Primary Stroke Centre
PUP: Point-of-Pickup
SÖSR: South-Eastern Healthcare Region
SUH: Sahlgrenska University Hospital
VGR: Västra Götaland Region
WGS 84: World Geodetic System 1984

## Author contributions

Concept and design: KT, GS, AR, KC, PA

Acquisition, analysis or interpretation of data: All authors

Drafting of the manuscript: KT

Critical review of the manuscript for important intellectual content: All authors

Final approval of the manuscript: All authors

Statistical analysis: KT, RG

Obtained funding: KT

Agreement on accountability for all aspects of the work: All authors

